# Machine Learning the Phenomenology of COVID-19 From Early Infection Dynamics

**DOI:** 10.1101/2020.03.17.20037309

**Authors:** Malik Magdon-Ismail

## Abstract

We present a robust data-driven machine learning analysis of the COVID-19 pandemic from its *early* infection dynamics, specifically infection counts over time. The goal is to extract actionable public health insights. These insights include the infectious force, the rate of a mild infection becoming serious, estimates for asymtomatic infections and predictions of new infections over time. We focus on USA data starting from the first confirmed infection on January 20 2020. Our methods reveal significant asymptomatic (hidden) infection, a lag of about 10 days, and we quantitatively confirm that the infectious force is strong with about a 0.14% transition from mild to serious infection. Our methods are efficient, robust and general, being agnostic to the specific virus and applicable to different populations or cohorts.

## 1 Introduction

As of March 1 2020, there was still much public debate on properties of the COVID-19 pandemic (see the CNN article, Cohen (2020)). For example, is asymptomatic spread of COVID-19 a major driver of the pandemic? There was no clear unimodal view, highlighting the need for robust tools to generate actionable quantitative intelligence on the nature of a pandemic from *early* and minimal data. One approach is scenario analysis. Recently, Chinazzi *et al*. (2020) used the Global Epidemic and Mobility Model (GLEAM) to perform infection scenario analyses on China COVID-19 data, using a networked meta-population model based on transportation hubs. A similar model for the US was reported in Wilson (2020) where the web-app predicted from 150,000 to 1.4 million infected cases by April 30, depending on the intervention level. Such scenario analysis requires user input such as infection sites and contagion-properties. However, a majority of infection sites may be hidden, especially if asymptomatic transmission is significant. Further, the contagion parameters are *unknown* and must be deduced, perhaps using domain expertise.

Data driven approaches are powerful. A long range regression analysis of COVID-19 out to 2025 on US data using immunological, epidemiological and seasonal effects is given in Kissler *et al*. (2020), which predicts recurrent outbreaks. We also follow a data-driven machine learning approach to understand early dynamics of COVID-19 on the first 54 days of US confirmed infection reports (downloadable from the European Center For Disease Control). We address the challenge of real-time data-intelligence from early data. Our approach is simple, requires minimal data or human input and generates actionable insights. For example, is asymptomatic spread significant? Our data-driven analysis says yes, emphatically. We even give quantitative estimates for the number of asymptomatic infections.

Early data is both a curse and a blessing. The curse is that “early” implies not much information, so quantitative models must be simple and robust to be identifiable from the data. The blessing is that early data is a peek at the pandemic as it roams free, unchecked by public health protocols, for to learn the true intentions of the lion, you must follow the beast on the savanna, not in the zoo. As we will see, much can be learned from early data and these insights early in the game, can be crucial to public health governance.

We analyzed daily confirmed COVID-19 cases from January 21 to March 14, the training or model calibration phase, the gray region in Figure 1. A more detailed plot of the model fit to the training data is in Figure 2. Qualitatively we see that the model captures the data, and it does so by setting four parameters:

**Figure 1:**
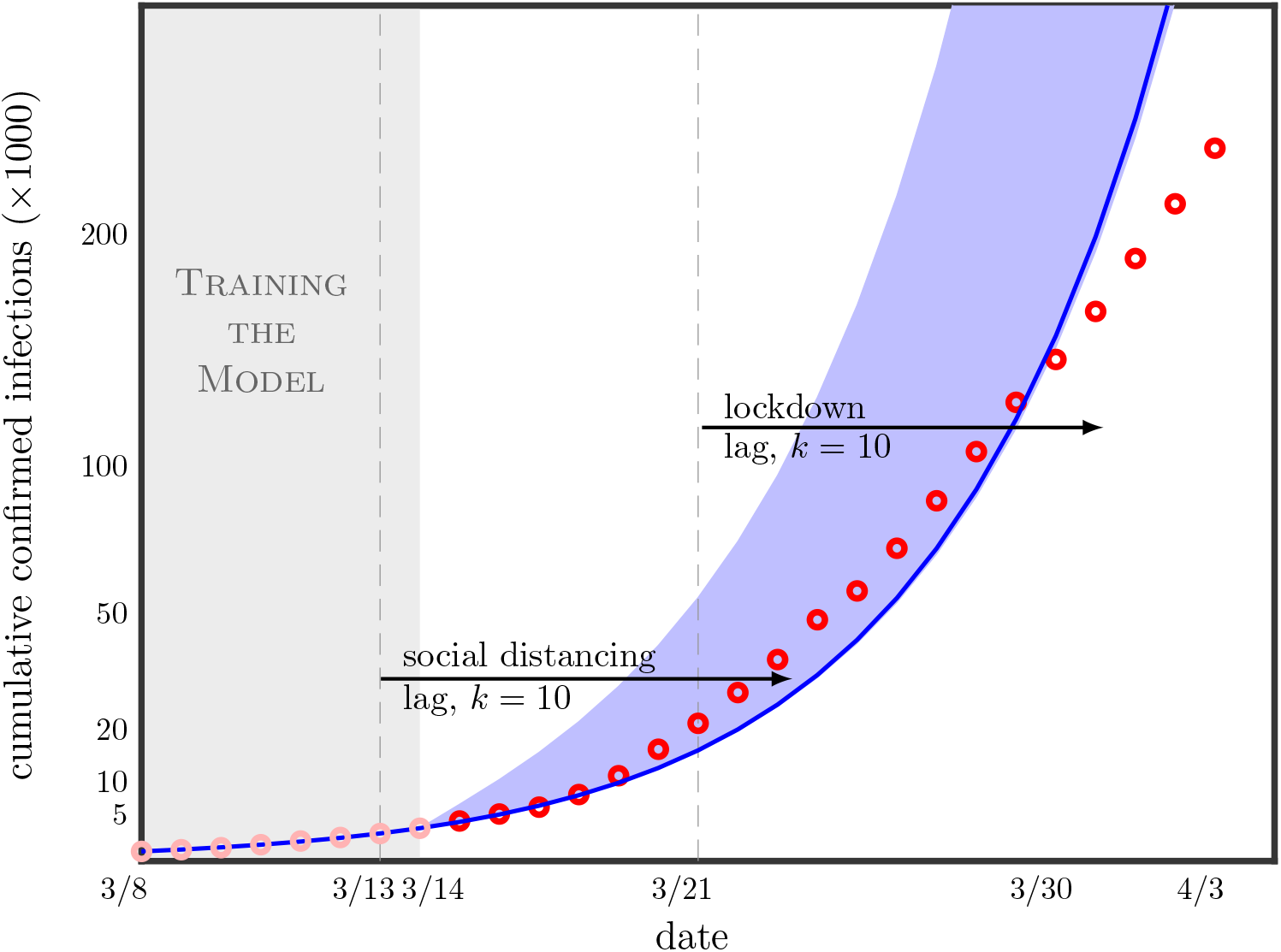
Training is the gray region and model predictions are the blue envelope. Observed infections fall away from predicteds, indicating that social distancing is working in agreement with our lag of 10 days (the two “kinks” in the curve). The figure emphasizes early data for learning about the pandemic, as later data is “contaminated” by public health protocols whose effects are hard to quantify. (Note: Dates are the time-stamps on the ECDC report (ECDC, 2020), which captures the previous day’s activity)

**Figure 2:**
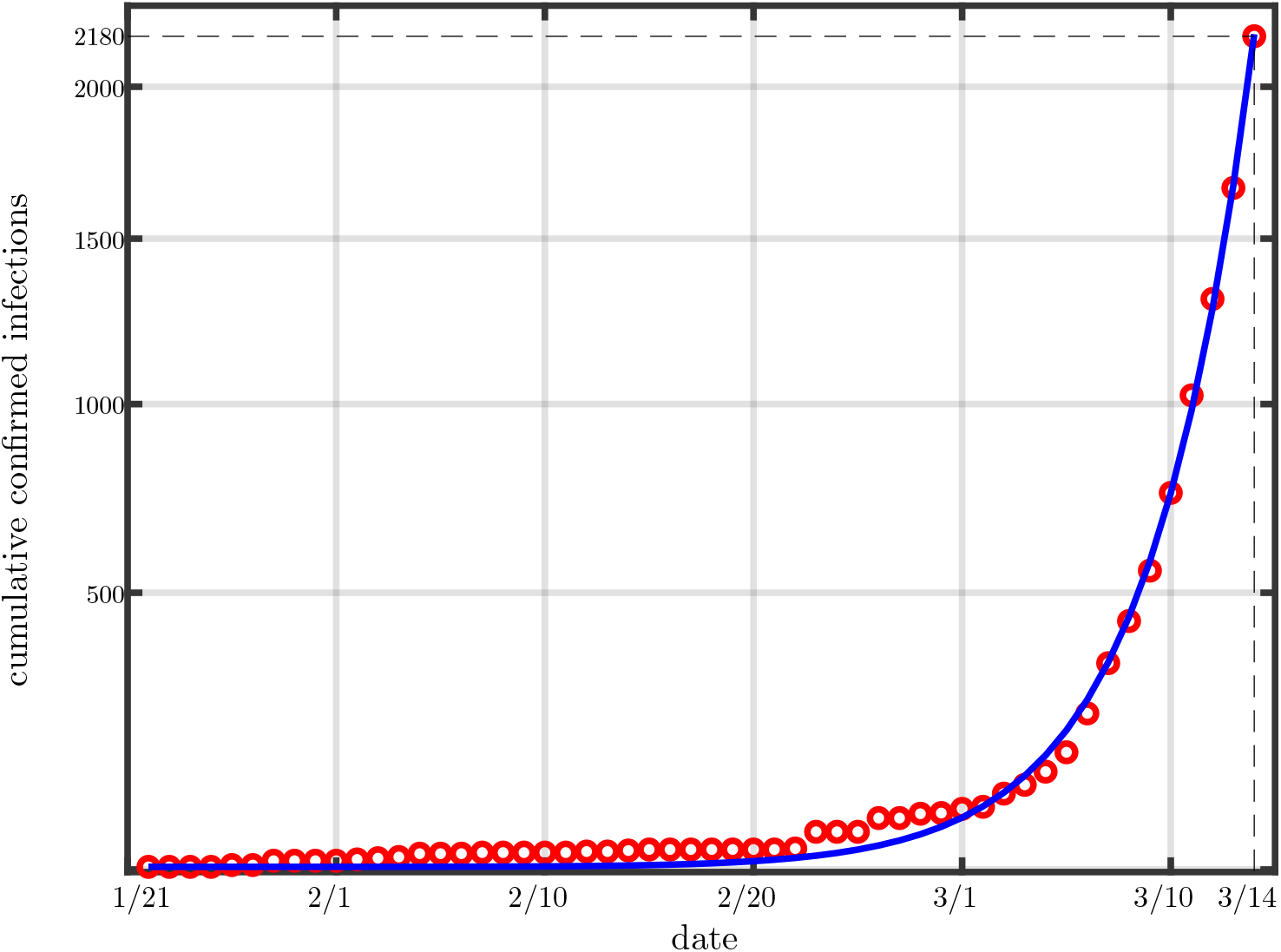
Model calibration to the early dynamics (first 54 infection counts) of (USA) COVID-19. Dates are the time-stamps on the ECDC report, which captures the previous day’s activity (e.g. the time stamp 1/21 is the infection on 1/20).

*β*, **asymptomatic** infectious force governing exponential spread

*γ*, virulence, the fraction of mild cases that become serious later

*k*, lag time for mild infection to become serious (an incubation time)

*M*_0_, Unconfirmed mild asymptomatic infections at time 0

Calibrating the model to the training data, gives the following information.

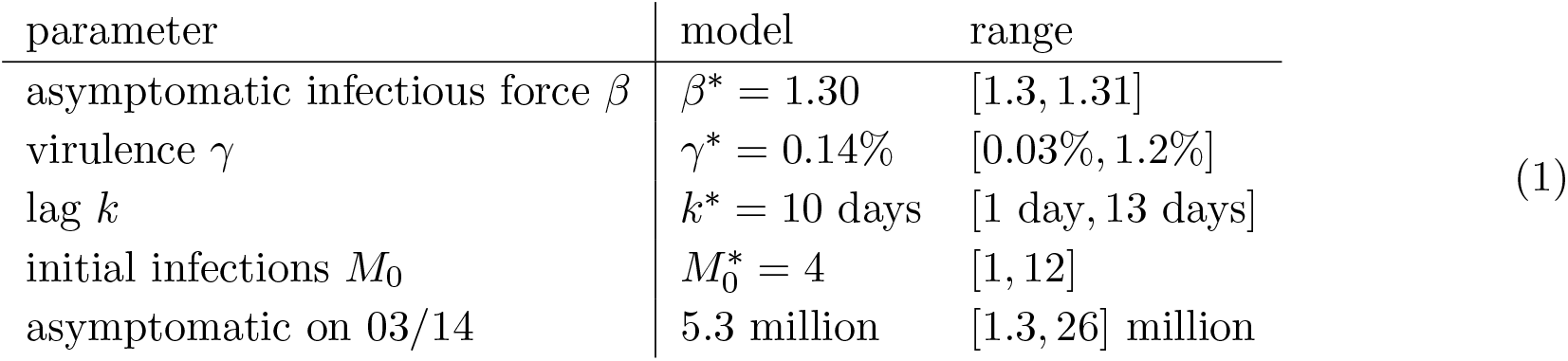

| parameter | model | range |
| --- | --- | --- |
| asymptomatic infectious force $\beta$ | $\beta^* = 1.30$ | [1.3, 1.31] |
| virulence $\gamma$ | $\gamma^* = 0.14\%$ | [0.03%, 1.2%] |
| lag $k$ | $k^* = 10$ days | [1 day, 13 days] |
| initial infections $M_0$ | $M_0^* = 4$ | [1, 12] |
| asymptomatic on 03/14 | 5.3 million | [1.3, 26] million |

The *asymptomatic* infectious force if left unchecked is 30% new cases per day (doubling in about days) and the virulence is *γ* = 0.14% (1 or 2 in a thousand conversions from mild infection to serious). Not all serious cases are fatal. The model output about 5.3 million asymptomatic cases as of 03/14 and a range from 1 to 26 million, a surprisingly high number. Such quantitative early intelligence has significance for public health protocols.

Beyond 03/14 in Figure 1 are the model predictions (blue envelope) and the red circles are the observed infection counts. How do we know the model predictions are honest, in that the red circles were in no way influenced the predictions. We are in a unique position to test the model because it is *time stamped* as version 1 of the preprint Magdon-Ismail (2020). The model has provably not changed since 03/14, and we just added test data as it arrived. The predictions in Figure 1 are in no way forward looking, data snooped or overfitted. We observe that the model and observed counts agree, modulo two “kinks” around 03/24 and 03/30, when the observed infections start falling away from the model. To understand the cause, the lag is important. Aggressive social distancing was implemented on about 03/13 and lockdowns around 03/21. A lag of *k* = 10 means the effects of these protocols will become apparent around 03/23 and 03/31 respectively.

The methods are general and can be applied to different cohorts. In Section 3.2 we do a cross-sectional country-based study. Our contributions are

- A methodology for quickly and robustly machine learning simple epidemiological models given coarse aggregated infection counts.
- Building a simple model with lag for learning from early pandemic dynamics.
- Application of the methodology within the context of COVID-19 to USA data. Our methods reveal significant asymptomatic (hidden) infection, a lag of about 10 days, and we quantitatively confirm that the infectious force is strong with about a 0.14% transition from mild to serious infection.
- Cross-sectional analysis of the pandemic dynamics across several countries.
- To our knowledge, the *only* tested predictions for COVID-19 due to our time-stamping of the predictions. Our results demonstrate the effectiveness of simple robust models for predicting pandemic dynamics from *early* data.

## 2 Model and Method

Our model is simple and robust. The majority of disease models aggregate individuals according to disease status, such as SI, SIR, SIS, Kermack and McKendrick (1927); Bailey (1957); Anderson and May (1992). We use a similar model by considering a mild infected state which can transition to a serious state. Early data allows us to make simplifying assumptions. In the early phase, when public health protocols have not kicked in, a confirmed infection is self-reported. That is, *you* decide to get tested. Why? Because the condition became serious. This is important. A confirmed case is a transition from mild infection to serious. This is not true later when, for example, public health protocols may institute randomized testing. At time *t* let there be *C*(*t*) confirmed cases and correspondingly *M* (*t*) mild unreported asymptomatic infections. The new confirmed cases at time *t* correspond to mild infections at some earlier time *t* − *k* which have now transitioned to serious and hence got self-reported. Let a fraction *γ* of those mild cases transitioned to serious,

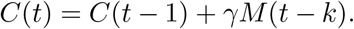

Another advantage of early dynamics is that we may approximate the growth from each infection as independent and exponential, according to the infectious force of the disease. So,

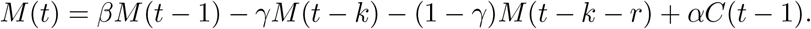

Here, the second term is the loss of mild cases that transitioned to serious, the third term is the remaining cases that don’t transition to serious recovering at some later time *r* and the fourth term accounts for new infections from confirmed cases. We will further simplify and assume that confirmed cases are fully quarantined and so *α* = 0 and recovery to a non-infectious state occurs late enough to not factor into early dynamics. Our simplified model is:

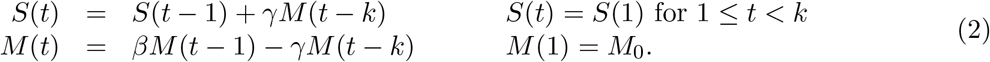

We set *t* = 1 at the first confirmed infection (Jan 21 in USA). Given *k, M*_0_, we get an approximate fit to the data by using a perturbation analysis to solve for *γ, β* that fit two points *S*(*τ* ) and *S*(*T* ):

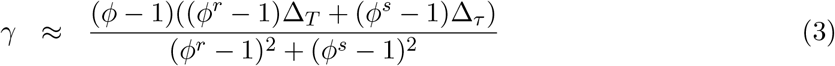

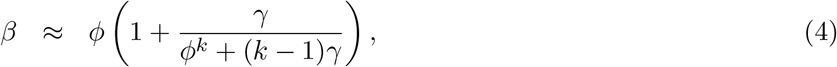

where,

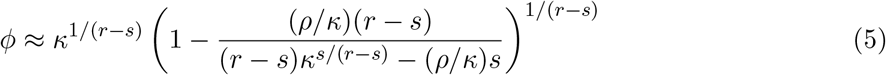

and *r* = *T* − *k, s* = *τ* − *k, κ* = (*S*(*T* ) − *S*(1))/(*S*(*τ* ) − *S*(1)) and *ρ* = *κ* − 1 (for details see the appendix). From this solution as a starting point, we can further optimize *γ, β* using a gradient descent which minimizes an error-measure that captures how well the parameters *β, γ, k, M*_0_ reproduce the observed dynamics in Figure 2, see for example Abu-Mostafa *et al*. (2012). We used a combination of root-mean-squared-error and root-mean-squared-percentage-error between observed dynamics and model predictions. By optimizing over *k, M*_0_, we obtain an optimal fit to the training data (Figure 2) using model parameters:

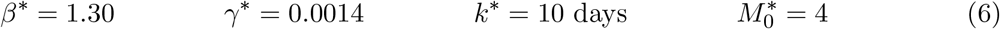

The asymptomatic infectious force, *β*, is very high, and corresponds to a doubling time of 2.6 days. The virulence at 0.14% seems comparable to a standard flu, though the virus may be affecting certain demographics much more severely than a flu. The incubation period of 10 days seems in line with physician observations. The data analysis predicts that when the first confirmed case appeared, there were 4 other infections in the USA. The parameters *β*^∗^, *γ*^∗^ and 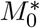 are new knowledge, gained with relative ease by calibrating a simple robust model to the early dynamics. But, these optimal parameters are not the whole story, especially when it comes to prediction.

The exhaustive search over *k, M*_0_, fixing *β* and *γ* to the optimal for that specific *k, M*_0_, produces several equivalent models We show the quality of fit for various (*k, M*_0_) in Figure 3(a). The deep-blue region contains essentially equivalent models within 0.5% of the optimal fit, our (user defined) error tolerance. The deep-blue region shows the extent to which the model is ill-identified by the data. Indeed, all these deep-blue models equally fit the data which results in a range of predictions. For robustness, we pick the white square in the middle of the deep-blue region, but note here that it is only one of the models which are consistent with the data. In making predictions, we should consider all equivalent models to get a range for the predictions that are all equally consistent with the data. Similarly, in Figure 3(b), we fix *k*^∗^ and 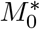 to their optimal robust values and show the uncertainty with respect to *β* and *γ* (the deep-blue region). Again, we pick the white square in the middle of the deep-blue region of equivalent models with respect to the data. Hence we arrive at our optimal parameters in Equation (6). By considering all models which are equally consistent with the data, we get the estimates toghether with the ranges in Equation 1. We emphasize that these error-ranges we report have nothing to do with the data, and are simply due to the ill-posedness of the inverse problem to inifer the model from finite data. Several models essentially fit the data equivalently. We do not include in our range the possible measurement errors in the data, although the two are related through the error tolerence used in defining “equivalent” models. More noise in the data would result in more models being treated as equivalent.

**Figure 3:**
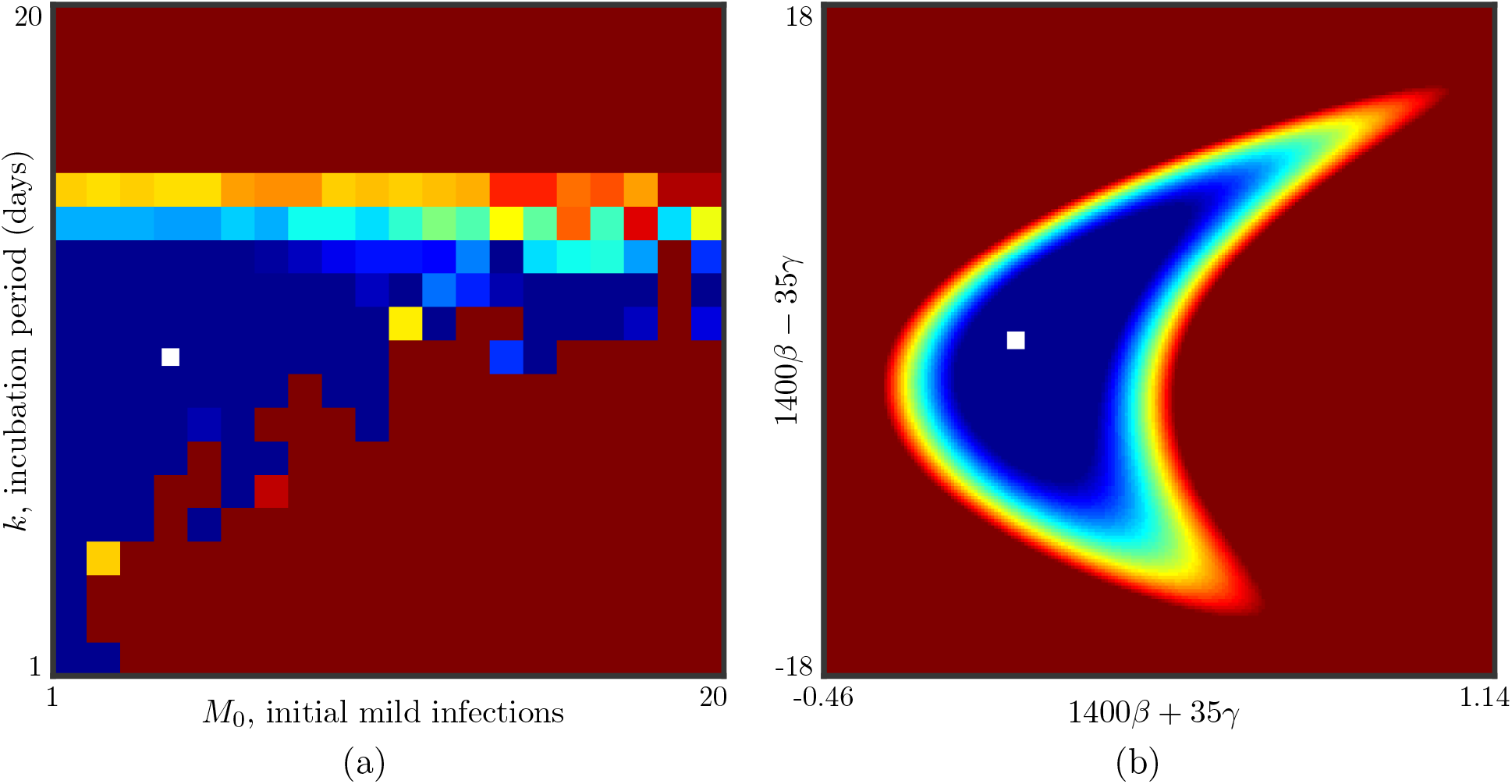
Uncertainty in model estimation. (a) Optimal fit-error over for different choices of *M*0 and *k*. Blue is better fit, red is worse. The deep-blue region corresponds to comparable models, within 0.5% of the optimal fit. The white square is the model chosen, 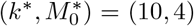 which has optimal fit and also is “robust” being in the middle of the deep-blue regions. (b) Model fit for the chosen *k*^∗^ and 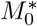. Again, the deep-blue is an equivalence class of optimal models. The “robust” model is the white square in the middle of the deep-blue region, (*β*^∗^, *γ*^∗^) = (1.30, 0.0014). The deep-blue regions represent uncertainty.

## 3 Results

As already mentioned, to get honest estimates, we must consider all models which are equally consistent with the data (deep-blue regions in Figure 3).

### 3.1 COVID-19 in USA

The model in Equation (1) gives the prediction of new infections in the table below. The cumulative predicted infections is the data plotted in Figure 1.

| Date | New Infections |  | Observed |
| --- | --- | --- | --- |
|  | Model Prediction | Range |  |
| March 15, 2020 | 665 | [657,2861] | **777** |
| March 16, 2020 | 866 | [856,3732] | **823** |
| March 17, 2020 | 1130 | [1114,4867] | **887** |
| March 18, 2020 | 1475 | [1450,6348] | **1766** |
| March 19, 2020 | 1924 | [1888,8280] | **2988** |
| March 20, 2020 | 2510 | [2459,10800] | **4835** |
| March 21, 2020 | 3275 | [3201,14088] | **5374** |
| March 22, 2020 | 4272 | [4168,18375] | **7123** |
| March 23, 2020 | 5574 | [5427,23968] | **8459** |
| March 24, 2020 | 7271 | [7067,31263] | **11236** |
| March 25, 2020 | 9487 | [9201,40789] | **8789** |
| March 26, 2020 | 12376 | [11980,53216] | **13693** |
| March 27, 2020 | 16146 | [15198,69430] | **16797** |
| March 28, 2020 | 21065 | [20309,90585] | **18695** |
| March 29, 2020 | 27482 | [26443,118180] | **19979** |
| March 30, 2020 | 35854 | [34430,154190] | **18360** |
| March 31, 2020 | 46777 | [44829,201170] | **21595** |
| April 1, 2020 | 61026 | [58369,262470] | **24998** |
| April 2, 2020 | 79617 | [75999,343440] | **27103** |

The predictions use the model in (1) and the ranges are obtained by using the space of models equally consistent with the data. March 15 and 16 data arrived at the time of writing and March 17 onward arrived after the time of writing time-stamped version 1 (Magdon-Ismail, 2020). Blue means in the range and red means outside the range.

### 3.2 Cross-Sectional Study By Countries

In the supplementary material we give details of our cross-sectional study across countries. The different countries have different cultures, social networks, mobility, demographics, as well as different times at which the first infection was reported (the “delay”). We calibrated independent models for each country and the resulting model parameters are in Table 1.

**Table 1:** Fit parameters for 20 countries.

| Country | $\beta$ | [min, max] | $\gamma$ | [min, max] | $k$ | [min, max] | $M_0$ | [min, max] |
| --- | --- | --- | --- | --- | --- | --- | --- | --- |
| Australia | 1.143 | [1.137,1.18] | 0.0047 | [0.0024,0.046] | 2 | [1,3] | 10 | [1,18] |
| Austria | 1.411 | [1.389,2.261] | 0.045 | [0.022,0.89] | 1 | [1,1] | 20 | [1,40] |
| Canada | 1.182 | [1.176,1.191] | 0.0014 | [0.00025,0.0099] | 3 | [1,3] | 10 | [1,40] |
| China | 1.312 | [1.297,1.332] | 5.18 | [3.75,8.26] | 15 | [14,16] | 6 | [5,9] |
| France | 1.297 | [1.293,1.301] | 0.05 | [0.0128,0.449] | 19 | [18,20] | 7 | [1,36] |
| Germany | 1.289 | [1.287,1.295] | 0.011 | [0.0039,0.044] | 8 | [1,8] | 4 | [1,10] |
| Iran | 1.596 | [1.563,1.709] | 0.78 | [0.71,0.79] | 4 | [3,4] | 30 | [29,40] |
| Ireland | 1.401 | [1.372,1.79] | 0.083 | [0.042,0.62] | 2 | [2,2] | 20 | [2,40] |
| Italy | 1.242 | [1.237,1.255] | 3.88 | [3.11,5.37] | 19 | [18,19] | 6 | [4,8] |
| Netherlands | 1.387 | [1.372,2.28] | 0.34 | [0.29,1.314] | 4 | [2,4] | 35 | [2,40] |
| Norway | 1.461 | [1.379,2.935] | 0.164 | [0.082,1.64] | 1 | [1,3] | 20 | [1,40] |
| Poland | 1.476 | [1.422,3.542] | 0.109 | [0.054,2.17] | 1 | [1,1] | 20 | [1,40] |
| Portugal | 1.4 | [1.356,3.077] | 0.0886 | [0.044,1.77] | 1 | [1,1] | 20 | [1,40] |
| South Africa | 1.563 | [1.513,2.65] | 0.057 | [0.031,1.15] | 1 | [1,2] | 20 | [1,37] |
| South Korea | 1.238 | [1.231,1.289] | 0.98 | [0.76,4.65] | 20 | [14,20] | 7 | [1,9] |
| Spain | 1.411 | [1.405,1.418] | 0.127 | [0.016,0.63] | 20 | [20,20] | 5 | [1,40] |
| Sweden | 1.356 | [1.343,1.372] | 0.064 | [0.0068,0.27] | 19 | [19,20] | 3 | [1,40] |
| Switzerland | 1.527 | [1.448,2.121] | 0.4 | [0.267,1.078] | 3 | [2,3] | 12 | [2,21] |
| UK | 1.252 | [1.248,1.257] | 0.68 | [0.412,0.878] | 19 | [17,20] | 1 | [1,1] |
| USA | 1.306 | [1.303,1.309] | 0.0014 | [0.0006,0.009] | 10 | [3,12] | 4 | [1,7] |

We primarily focused on the infectious force *β*, which has significant variability, and we studied how *β* statistically depends on a number of country-specific parameters factors. In the supplementary material, we give details of the study and the quantitative results. Qualitatively, we find:

- A larger delay in the virus reaching a country indicates a larger *β*. The more that has been witnessed, the faster the spread. That seems unusual, but is strongly indicated. We do not have a good explanation for this effect. It could be an artefact of testing procedures not being streamlined, so early adopters of the pandamic presenting as serious were not detected.
- Population density at the infection site has a strong positive effect but the country’s population density does not.
- There is faster spread in countries with more people under the poverty level defined as the percentage of people living on less than $5.50 per day.
- Median age has a strong effect. Spread is faster in younger populations. The youth are more mobile and perhaps also more carefree.
- Wealth and per-capita income have a slight negative effect. Spread is slower in richer countries, perhaps due to risk-aversion, higher levels of education and less reliance on public transportation. Whatever the cause, it does have an impact, but relatively smaller than the other effects.

## 4 Conclusion

Early dynamics allows us to learn useful properties of the pandemic. Later dynamics may be contaminated by human intervention, which renders the data less useful without a more general model. We learned a lot from the early dynamics of COVID-19. It’s infectious force, virulence, incubation period, unseen infections and predictions of new confirmed cases. All this, albeit within error tolerances, from a simple model and little data. Asymptomatic infection is strong, around 30%, converting to serious at a rate at most 1.2%. There is significant uncertainty in the lag, from 1 up to 13 days, and we estimate 5.3 million asymptomatic infections as of 03/14, the range being from 1 to 26 million. Such information is useful for planning and health-system preparedness. Are our parameters correct? We were in a unique position to *test* our predictions because our model was *time-stamped* as version 1 of the preprint Magdon-Ismail (2020).

A side benefit of the model predictions is as a benchmark against which to evaluate public health interventions. If moving forward, observed new infections are low compared to the data in, it means the interventions are working by most likely reducing *β*. Starting on about March 25, the observed infections starts falling off and we observe a flattening by March 28. The US instuted broad and aggressive social distancing protocols starting on or before March 13 and even stronger lockdown around March 21, which is consistent with the data and the model’s lag of *k* = 10. Without such quantitative targets to compare with, it would be hard to evaluate intervention protocols.

Our approach is simple and works with coarse, aggregated data. But, there are limitations.

- The independent evolution of infection sites only applies to early dynamics. Hence, when the model infections increase beyond some point, and the pandemic begins to saturate the population, a more sophisticated network model that captures the dependence between infection sites would be needed Balcan *et al*. (2009); Hill *et al*. (2010); Salathé and Jones (2010); Keeling and Eames (2005); Chinazzi *et al*. (2020).
- While we did present an optimal model, it should be taken with a grain of salt because many models are nearly equivalent, resulting in prediction uncertainty.
- The model and the interpretation of its parameters will change once public health protocols kick in. The model may have to be re-calibrated (for example if *β* decreases) and the parameters may have to be reinterpreted (for example *γ* is a virulence only in the self-reporting setting, not for random testing). It is also possible to build a more general model with an early phase *β*_1_ and a latter phase *β*_2_ (after social distancing). But, beware, for a more general sophisticated model looks good *a priori* until it comes time to calibrate it to data, at which point it becomes unidentifiable.
- The model was learned on USA data. The learned model parameters may not be appropriate for another society. The virulence could be globally invariant, but it could also depend on genetic and demographic factors like age, as well as what “serious” means for the culture - that is when do you get yourself checked. In a high-strung society, you expect high virulence-parameter since the threshold for serious is lower. One certainly expects the infectious force to depend on the underlying social network and interaction patterns between people, which can vary drastically from one society to another and depending on interventions. Hence, one should calibrate the model to country specific data to gain more useful insights.

### The Lag, k, and Public Policy

The lag *k* is important for public policy due to how public policy can be driven by human psychology. The Human tendency is to associate any improvement in outcome to recent actions. However, if there is a lag, one might prematurely reward those recent actions instead of the earlier actions whose effects are actually being seen. Such lags are present in traditional machine learning, for example the delayed reward in reinforcement learning settings. Credit assignment to prior actions in the face of delayed reward is a notoriously difficult task, and this remains so with humans in the loop. Knowledge of the lag helps to assign credit appropriately to prior actions, and the public health setting is no exception.

## Data Availability

All data is publicly available.

## Acknowledgments

We thank Abhijit Patel, Sibel Adali and Zainab Magdon-Ismail for provoking discussions on this topic.

## A Fitting The Model

Recall the model,

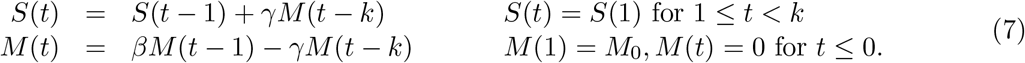

For fixed *k, M*_0_, we must perform a gradient descent to optimize *β, γ*. Unfortunately, the dependence on *β* is exponential and hence very sensitive. So if the starting point is not chosen carefully, the optimization gets stuck in a very flat region, and many millions of iterations are needed to converge. Hence it is prudent to choose the starting conditions carefully. To do so, we need to analyze the recursion. First, we observe that the recursion for *M* (*t*) is a standalone *k*-th order recurrence. For 1 ≤ *t* ≤ *k, M* (*t*) = *M*_0_*β*^*t*−1^, hence, we can guess a solution *M* (*t*) = *M*_0_*β*^*k*−1^*ϕ*^*t*−*k*^, for *t* > *k*, which requires

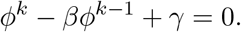

We do a perturbation analysis in *γ →* 0. At *γ* = 0, *ϕ* = *β*, so we set *ϕ* = *β* + *ϵ*, to get

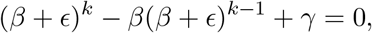

which to first order in *ϥ* is solved by *ϥ* ≈ −*γ*/*β*^*k*−1^ and so

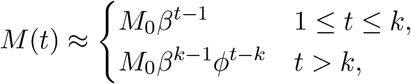

where *ϕ* = *β*(1 − *γ*/*β*^*k*^). Given this approximation, we can solve for *S*(*t*),

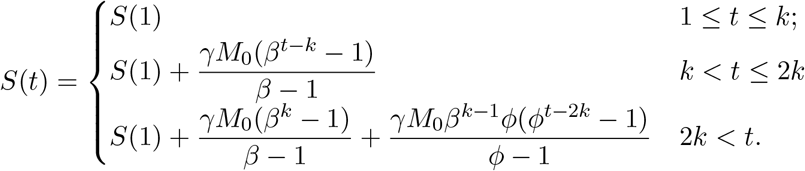

Since *ϕ* = *β* + *O*(*γ*), for *t* > 2*k*, we can approximate *S*(*t*) as,

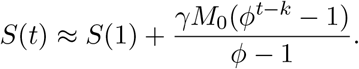

We can independently control two parameters *ϕ* and *γ*. We use this to match the observed *S*(*t*) at two time points. Since the growth is exponential, we match the end time, *S*(*T* ) and some time *τ* in the middle, for example *τ* = 3*T*/4 . Let Δ_*T*_ = (*S*(*T* ) − *S*(1))/*M*_0_ and Δ_*τ*_ = (*S*(*τ* ) − *S*(1))/*M*_0_. Then,

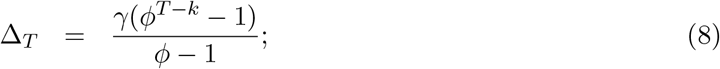

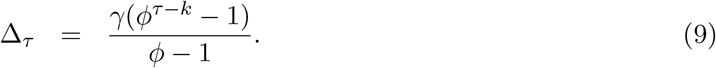

Dividing gives Δ_*T*_ /Δ_*τ*_ = (*ϕ*^*T* −*k*^ − 1)/(*ϕ*^*τ*−*k*^ − 1) ≈ *ϕ*^*T* −*τ*^, because *ϕ* > 1. Let us consider the equation *κ* = (*ϕ*^*r*^ − 1)/(*ϕ*^*s*^ − 1), which gives *ϕ*^*r*^ − *κϕ*^*s*^ + *κ* − 1 = 0, or more generally *ϕ*^*r*^ − *κϕ*^*s*^ + *ρ* = 0, where *r* > *s* > 1 and *κ* > *ρ* ≫ 1. This means *ϕ* > 1. When *ρ* = 0, we have *ϕ* = *κ*, so we do a perturbation analysis with *ϕ*^*r*−*s*^ = *κ* + *ϵ*, and our perturbation parameter is *ϵ*. Then, *ϕ*^*r*^ = (*κ* + *ϵ*)*ϕ*^*s*^ and plugging into the equation gives

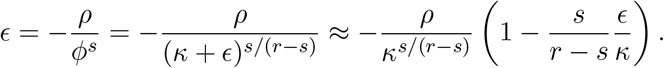

Solving for *ϥ* gives *ϥ* ≈ −*ρ*(*r* − *s*)/((*r* − *s*)*κ*^*s*/(*r*−*s*)^ + *s*), which gives

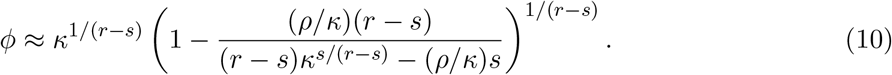

For our setting, *r* = *T* − *k, s* = *τ* − *k, κ* = Δ_*T*_ /Δ_*τ*_ and *ρ* = *κ* − 1. Finally, since *ϕ* is approximate, we may not be able to satisfy both equations in (9), hence we can instead minimize the mean squared error, which gives

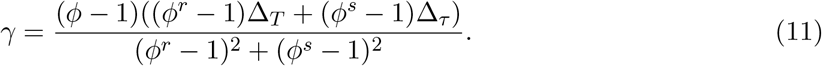

We now need to get *β* which satisfies *ϕ* = *β*(1 − *γ*/*β*^*k*^). Again, we do a perturbation analysis, omitting the details, to obtain

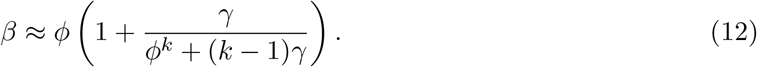

If one wishes, a fixed point iteration starting at the above will quickly approach a solution to *ϕ* = *β*(1 − *γ*/*β*^*k*^).

We show the approximate fit on the US data (Figure 4). We show the optimal fit, the initial fit using the parameters constructed from (12) and (11). The parameters and fit error are

| | $\beta$ | $\gamma$ | fit error |
| --- | --- | --- | --- |
| optimal | 1.306 | 0.0013282 | 1.6521 |
| Equations (12) and (11) | 1.3055 | 0.0013423 | 1.6619 |

**Figure 4:**
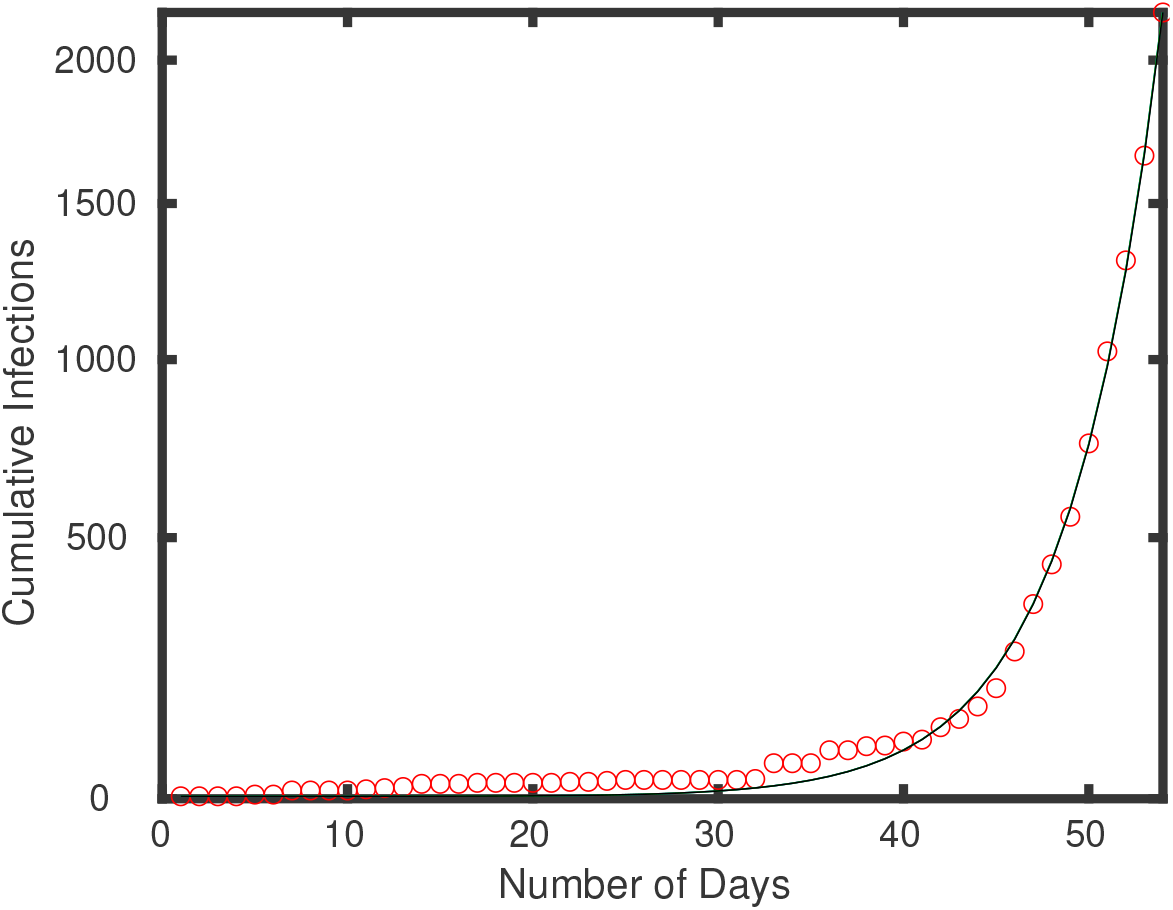
Approximate fit to USA data is essentially on top of optimal fit.

The approximate fit works pretty well, and is certainly good enough to initialize an optimization. Note that to get an even better starting point for the gradient descent optimization, assuming *T, τ* ≥ 2*k*, one could simultaneously solve the three equations

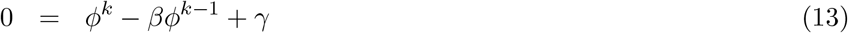

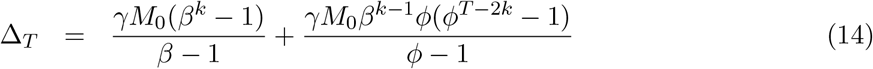

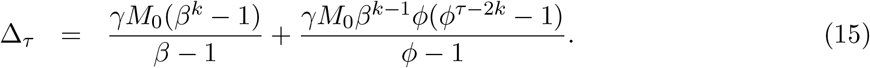

## B Cross-Sectional Country Study

We perform our analysis on early dynamics data available from the ECDC giving infection numbers starting from December 31, 2019 ECDC (2020). We use data for 20 countries selected qualitatively because they appear to have reasonably efficient testing procedures for self-reported cases. We iniclude China data for completeness, even though China dynamics since December 31 are not early dynamics. We show these countries below, together with some demographic data which might determine spread dynamics: Start City; Start and End dates; Delay to first infection in days; Population Density; Median Age (Wikipedia, 2020); Wealth as defined by adjusted net national income per capita (World-Bank, 2017); Average Income (World-Data-Info, 2015); Poverty Level (Wikipedia, Source: World Bank, 2020) defined by living on less than $5.50 per day; Population Density around the first infection site for the country.

From the public health perspective, perhaps the most important parameter is *β*, since actions can be taken to mitigate the spread by reducing *β*, whereas *γ, k* and *M*_0_ are somewhat givens for the country. We show the fits in Table 3. As you can see, there is much variability in *β*.

**Table 3:** Fit parameters for 20 countries.

| Country | $\beta$ | [min, max] | $\gamma$ | [min, max] | $k$ | [min, max] | $M_0$ | [min, max] |
| --- | --- | --- | --- | --- | --- | --- | --- | --- |
| Australia | 1.143 | [1.137,1.18] | 0.0047 | [0.0024,0.046] | 2 | [1,3] | 10 | [1,18] |
| Austria | 1.411 | [1.389,2.261] | 0.045 | [0.022,0.89] | 1 | [1,1] | 20 | [1,40] |
| Canada | 1.182 | [1.176,1.191] | 0.0014 | [0.00025,0.0099] | 3 | [1,3] | 10 | [1,40] |
| China | 1.312 | [1.297,1.332] | 5.18 | [3.75,8.26] | 15 | [14,16] | 6 | [5,9] |
| France | 1.297 | [1.293,1.301] | 0.05 | [0.0128,0.449] | 19 | [18,20] | 7 | [1,36] |
| Germany | 1.289 | [1.287,1.295] | 0.011 | [0.0039,0.044] | 8 | [1,8] | 4 | [1,10] |
| Iran | 1.596 | [1.563,1.709] | 0.78 | [0.71,0.79] | 4 | [3,4] | 30 | [29,40] |
| Ireland | 1.401 | [1.372,1.79] | 0.083 | [0.042,0.62] | 2 | [2,2] | 20 | [2,40] |
| Italy | 1.242 | [1.237,1.255] | 3.88 | [3.11,5.37] | 19 | [18,19] | 6 | [4,8] |
| Netherlands | 1.387 | [1.372,2.28] | 0.34 | [0.29,1.314] | 4 | [2,4] | 35 | [2,40] |
| Norway | 1.461 | [1.379,2.935] | 0.164 | [0.082,1.64] | 1 | [1,3] | 20 | [1,40] |
| Poland | 1.476 | [1.422,3.542] | 0.109 | [0.054,2.17] | 1 | [1,1] | 20 | [1,40] |
| Portugal | 1.400 | [1.356,3.077] | 0.0886 | [0.044,1.77] | 1 | [1,1] | 20 | [1,40] |
| South Africa | 1.563 | [1.513,2.65] | 0.057 | [0.031,1.15] | 1 | [1,2] | 20 | [1,37] |
| South Korea | 1.238 | [1.231,1.289] | 0.98 | [0.76,4.65] | 20 | [14,20] | 7 | [1,9] |
| Spain | 1.411 | [1.405,1.418] | 0.127 | [0.016,0.63] | 20 | [20,20] | 5 | [1,40] |
| Sweden | 1.356 | [1.343,1.372] | 0.064 | [0.0068,0.27] | 19 | [19,20] | 3 | [1,40] |
| Switzerland | 1.527 | [1.448,2.121] | 0.4 | [0.267,1.078] | 3 | [2,3] | 12 | [2,21] |
| UK | 1.252 | [1.248,1.257] | 0.68 | [0.412,0.878] | 19 | [17,20] | 1 | [1,1] |
| USA | 1.306 | [1.303,1.309] | 0.0014 | [0.0006,0.009] | 10 | [3,12] | 4 | [1,7] |

### B.1 Explaining *β*

We perform a simple statistical analysis to test if *β* can be explained by any of the country parameters in Table 2. We include the delay as a global explanatory variable, which would account for a global increase in vigilence as time passes and awareness of the pandemic increases. One expects *β* to decrease with the delay. A table of correlations of *β* with the various parameters is shown below. For our analysis we use the best case *β*, although similar results follow from the optimal *β*.

**Table 2:** Comparison of countries used in the study.

| Country | Code | Start City | Start | End | Delay | Den | Age | Wealth | Income | Pov. | Den-Init |
| --- | --- | --- | --- | --- | --- | --- | --- | --- | --- | --- | --- |
| Australia | AU | Melb./Syd. | 1/25 | 3/14 | 26 | 3 | 38.7 | 41489 | 53230 | 1.2 | 425 |
| Austria | AT | Innsbr./Vien. | 2/26 | 3/14 | 58 | 106 | 44 | 38748 | 49310 | 0.9 | 3000 |
| Canada | CA | Toronto | 1/26 | 3/14 | 27 | 4 | 42.2 | 36802 | 44940 | 1 | 4150 |
| China | CN | Wuhan | 12/31 | 2/5 | 1 | 145 | 37.4 | 6568 | 9460 | 27.2 | 1200 |
| France | FR | Bordeaux | 1/25 | 3/14 | 26 | 123 | 41.4 | 32672 | 41080 | 0.2 | 5000 |
| Germany | DE | Coppingen | 1/28 | 3/14 | 29 | 233 | 47.1 | 37791 | 47090 | 0.2 | 972 |
| Iran | IR | Qom/Tehran | 2/20 | 3/14 | 52 | 54 | 30.3 | 4238 | 5470 | 11.6 | 15500 |
| Ireland | IE | Dublin | 3/3 | 3/14 | 64 | 70 | 36.8 | 37988 | 61390 | 0.7 | 4588 |
| Italy | IT | Rome | 1/31 | 3/14 | 32 | 200 | 45.5 | 26537 | 33730 | 3.5 | 2232 |
| Netherl. | NL | Tilburg | 2/28 | 3/14 | 60 | 420 | 42.6 | 40545 | 51260 | 0.5 | 1852 |
| Norway | NO | Tr. Finn/Oslo | 2/27 | 3/14 | 59 | 17 | 39.2 | 61865 | 80610 | 0.2 | 1400 |
| Poland | PL | Zielona Gora | 3/6 | 3/14 | 67 | 123 | 40.7 | 11650 | 14100 | 2.1 | 504 |
| Portugal | PT | Porto | 3/3 | 3/14 | 64 | 112 | 42.2 | 17188 | 21990 | 3 | 6900 |
| S. Africa | ZA | Gaut./Dur. | 3/8 | 3/14 | 69 | 48 | 27.1 | 4942 | 5750 | 57.1 | 4000 |
| S. Korea | KR | Daegu | 1/20 | 3/4 | 21 | 517 | 41.8 | 24028 | 30600 | 1.2 | 2818 |
| Spain | ES | La Gom/Ten. | 2/1 | 3/14 | 33 | 93 | 42.7 | 23216 | 29340 | 2.9 | 250 |
| Sweden | SE | Jonkoping | 2/1 | 3/14 | 33 | 23 | 41.2 | 45149 | 55490 | 1 | 2100 |
| Switzer. | CH | Tin./Bas./Zur. | 2/26 | 3/14 | 58 | 208 | 42.4 | 64307 | 84410 | 0 | 6000 |
| UK | UK | Newcas.Tyme | 1/31 | 3/14 | 32 | 274 | 40.5 | 34171 | 41770 | 0.7 | 233 |
| USA | US | Seat./Snohom. | 1/21 | 3/14 | 22 | 34 | 38.1 | 51485 | 63080 | 2 | 3430 |

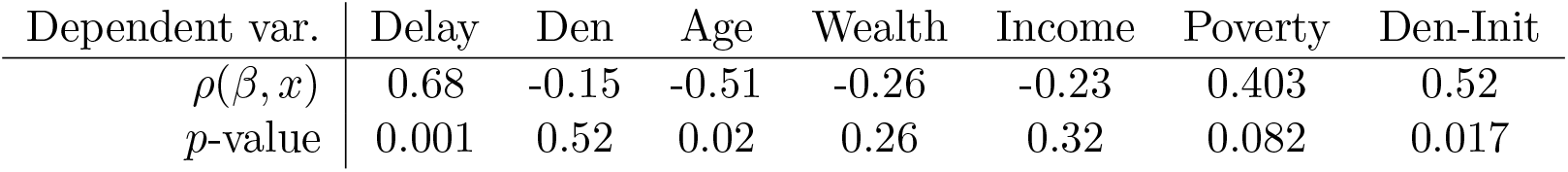

| Dependent var. | Delay | Den | Age | Wealth | Income | Poverty | Den-Init |
| --- | --- | --- | --- | --- | --- | --- | --- |
| $\rho(\beta, x)$ | 0.68 | -0.15 | -0.51 | -0.26 | -0.23 | 0.403 | 0.52 |
| $p$-value | 0.001 | 0.52 | 0.02 | 0.26 | 0.32 | 0.082 | 0.017 |

As expected, there is a very significant correlation of *β* with delay, but in the opposite direction.

- The larger the delay, the larger is *β*. The more a country has observed, the faster the spread in that country. That seems unusual but seems strongly indicated by the data.
- Population density at the infection site has a strong positive effect but the country’s population density does not.
- There is faster spread in poorer countries.
- Median age has a strong effect. Spread is faster in younger countries. The youth are more mobile and perhaps also more carefree.
- There is a slight negative effect from wealth and per-capita income. Spread is slower in richer countries. Perhaps this is due to more risk-aversion, perhaps higher levels of education, perhaps less use of public transportation. Whatever the cause, it does have an impact, but relatively smaller than the other effects.

We now use regularized regression to perform a linear model fit to explain *β*. To make the weight magnitudes meaningful, we normalize the data. We use a leave-one-out cross validation to select the optimal regularization parameter (which happens to be 20). The optimal regularized fit with this regularization parameter gives a new feature

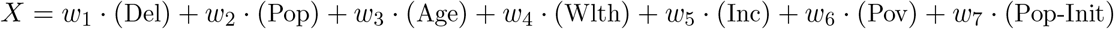

The learned weights and the their ranges which yield a cross-validation error within 10% of optimal are shown in the table.

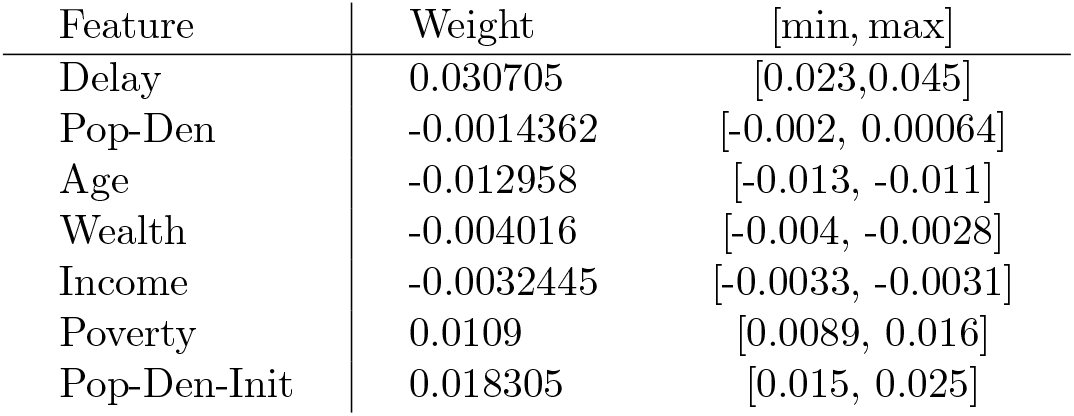

The predictions of *β* using this feature are also shown in Figure 5. A statistical regression model using these data produces the fit:

| Feature | Estimate | SE | *t*-Stat | *p*-Value |
| --- | --- | --- | --- | --- |
| (Intercept) | 1.2213 | 0.32759 | 3.7282 | 0.0028836 |
| Delay | 0.0030891 | 0.0010042 | 3.0762 | 0.0096064 |
| Pop-Den | 2.0281e-05 | 0.00014317 | 0.14166 | 0.8897 |
| Age | -0.0013599 | 0.0074969 | -0.1814 | 0.85908 |
| Wealth | 2.274e-06 | 7.9597e-06 | 0.28569 | 0.77999 |
| Income | -1.8495e-06 | 6.0964e-06 | -0.30338 | 0.7668 |
| Poverty | 0.0019366 | 0.0024657 | 0.7854 | 0.44745 |
| Pop-Den-Init | 9.6516e-06 | 6.6452e-06 | 1.4524 | 0.17203 |

**Figure 5:**
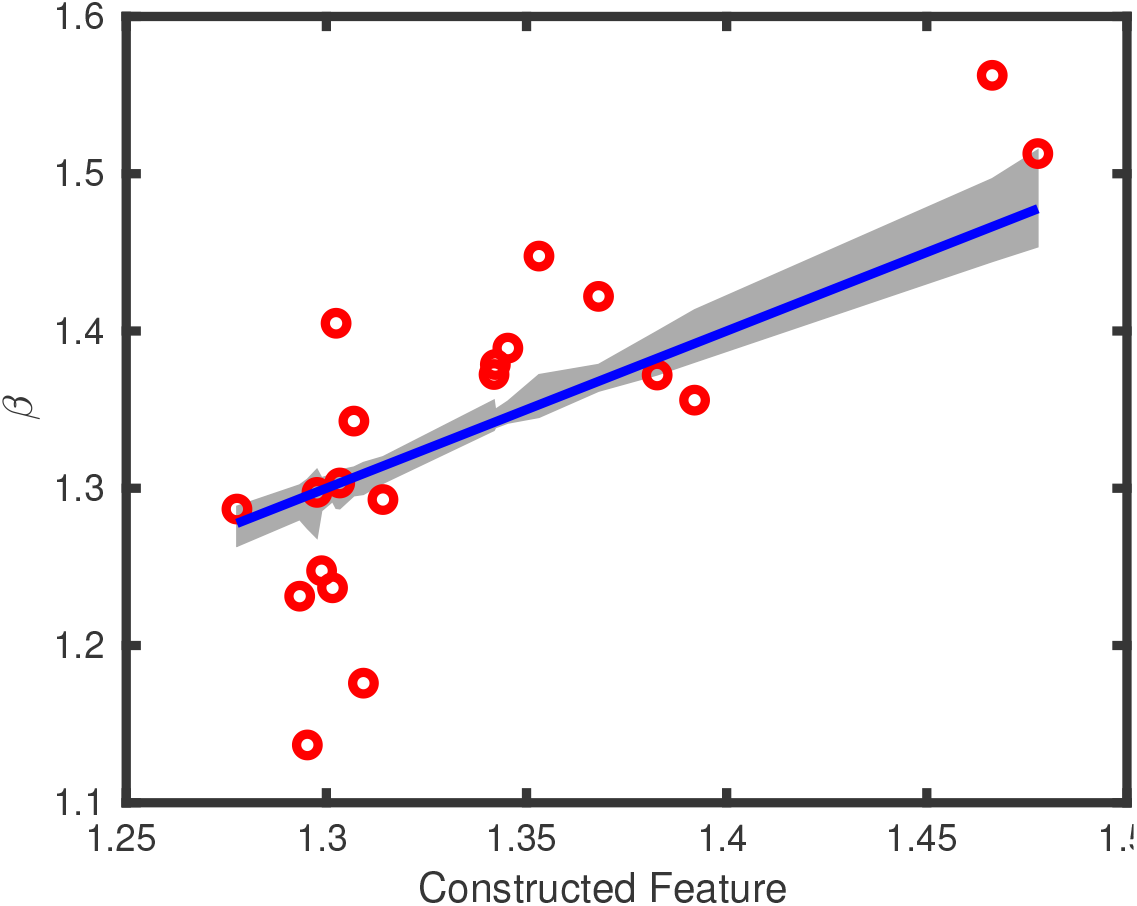
Optimal feature to predict *β*, within a cross-valudation setting to select the regularization parameter. The gray region is the range of the predicted value for each country. The *R*^2^ = 0.57, so the cross-validation based optimal linear feature captures 57% of the residual variance.

The statistical regression model also identifies positive weights on delay, population density at the initial site and poverty in that order of significance.

As we observed from the correlations, Delay, Poverty and Population Density at the initial infection site have strong positive weights. Age has a strong negative weight. Wealth and income have weak negative effects, but non-zero. The population density of the country as a whole seems to have no effect, with a weight range that includes 0.

## Notes

### Competing Interest Statement

The authors have declared no competing interest.

### Funding Statement

None

## References

Abu-Mostafa, Y., Magdon-Ismail, M., and Lin, H.-T. (2012). Learning From Data: A Short Course. aml-book.com.

Anderson, R. and May, R. (1992). Infectious diseases of humans. Oxford University Press.

Bailey, N. (1957). The mathematical theory of epidemics. Griffin.

Balcan, D., Colizza, V., Gonçalves, B., Hu, H., Ramasco, J. J., and Vespignani, A. (2009). Multiscale mobility networks and the spatial spreading of infectious diseases. Proceedings of the National Academy of Sciences, 106(51), 21484–21489.

Chinazzi, M., Davis, J. T., Ajelli, M., Gioannini, C., Litvinova, M., Merler, S. y Piontti, A. P., Mu, K., Rossi, L., Sun, K., Viboud, C., Xiong, X., Yu, H., Halloran, M. E., Jr., I. M. L., and Vespignani, A. (2020). The effect of travel restrictions on the spread of the 2019 novel coronavirus (covid-19) outbreak. Science, page DOI: 10.1126/science.aba9757.

Cohen, E. (2020). Infected people without symptoms might be driving the spread of coronavirus more than we realized. https://www.cnn.com. (Elizabeth Cohen is Senior Medical Correspondent for CNN).

ECDC (2020). Geographic distribution of covid-19 cases worldwide. https://www.ecdc.europa.eu. (European Center for Disease Control).

Hill, A. L., Rand, D. G., Nowak, M. A., and Christakis, N. A. (2010). Infectious disease modeling of social contagion in networks. PLOS Computational Biology, 6(11).

Keeling, M. J. and Eames, K. T. (2005). Networks and epidemic models. Journal of the Royal Society Interface.

Kermack, W. and McKendrick, A. (1927). A contribution to the mathematical theory of epidemics. Proceedings of the Royal Society A, pages 700–721.

Kissler, S. M., Tedijanto, C., Goldstein, E. M., Grad, Y. H., and Lipsitch, M. (2020). Projecting the transmission dynamics of sars-cov-2 through the post-pandemic period. medRxiv preprint, Harvard.

Magdon-Ismail, M. (2020). Machine learning the phenomenology of covid-19 from early infection dynamics. arXiv preprint, https://arxiv.org/abs/2003.07602.

Salathé, M. and Jones, J. H. (2010). Dynamics and control of diseases in networks with community structure. PLOS Computational Biology, 6(4).

Wikipedia (2020). www.wikipedia.com.

Wikipedia, Source: World Bank (2020). List of countries by percentage of population living in poverty. www.wikipedia.com.

Wilson, C. (2020). Exclusive: Here’s how fast the coronavirus could infect over 1 million americans. TIME web-applet available for scenario analysis.

World-Bank (2017). Adjusted net national income per capita (current us$). https://data.worldbank.org.

World-Data-Info (2015). Average income around the world. https://data.worlddata.info.

